# Performance of rapid diagnostic tests, microscopy, and qPCR for detection of parasites among community members with or without symptoms of malaria in villages with high levels of artemisinin partial resistance in North-western Tanzania

**DOI:** 10.1101/2024.09.30.24314608

**Authors:** Rule Budodo, Catherine Bakari, Salehe S. Mandai, Misago D. Seth, Filbert Francis, Gervas A. Chacha, Angelina J. Kisambale, Daniel P. Challe, Daniel A. Petro, Dativa Pereus, Rashid A. Madebe, Ruth B. Mbwambo, Ramadhani Moshi, Sijenunu Aaron, Daniel Mbwambo, Abdallah Lusasi, Stella Kajange, Samwel Lazaro, Ntuli Kapologwe, Celine I. Mandara, Deus S. Ishengoma

## Abstract

**Background:** Despite the implementation of different control interventions, infections in the communities (among asymptomatic and symptomatic individuals) still play a crucial role in sustaining malaria transmission. This study evaluated the performance of rapid diagnostic tests (RDTs), microscopy, and quantitative PCR (qPCR) in detecting malaria parasites among community members in five villages of Kyerwa district, Kagera region, an area where artemisinin partial resistance (ART-R) has been recently confirmed.

**Methods:** A community cross-sectional survey of asymptomatic and symptomatic participants (n=4454) aged ≥6 months was conducted in July and August 2023. Malaria infections were detected using RDTs, microscopy, and qPCR (using 18S RNA gene). Performance of RDTs and microscopy was assessed by sensitivity, specificity, and predictive values, using qPCR as the reference method. Factors affecting the accuracy of these methods were explored using a multivariate logistic regression model.

**Results:** The prevalence of malaria infections was 44.4% (n = 1979/4454), 32.1% (n = 1431/4454), and 39.8% (n = 1771/4454) by RDTs, microscopy, and qPCR, respectively. The prevalence of *P. malariae* and *P. ovale* mono-infection by microscopy was 0.2% (n = 7/4454) and 0.3% (n = 12/4454), while by qPCR was 0.4% (n = 16/4454) and 0.5% (n = 24/4454), respectively. The geometric mean parasite densities (GMPDs) by microscopy were 642 (95% confidence intervals (CI) = 570 - 723), 126 (95% CI = 98 - 162), and 124 (95% CI = 82 - 160) asexual parasites/µL; while by qPCR, the GMPDs were 1180 (95% CI = 1032 - 1349), 44 (95% CI = 32 - 61), and 50 (95% CI = 29 - 89) parasites/µL for *P. falciparum, P. ovale spp*, and *P. malariae*, respectively. The sensitivity and specificity of RDTs were 94.0% (95% CI = 92.8% - 95.1%) and 87.5% (95% CI = 86.2% - 88.7%), respectively; whereas those of microscopy were 74.6% (95% CI = 72.5% - 76.6%) and 95.2% (95% CI = 94.3% - 96.0%), respectively. The sensitivity of RDTs, and microscopy was low at very low parasitaemia (<100 parasite/μL), but increased significantly with increasing parasitaemia, reaching ≥99.6% at >10000 parasites/μL (p<0.001).

**Conclusion:** Higher prevalence of malaria was detected and the performance of RDTs and qPCR were comparable, but microscopy had lower performance. Higher sensitivity by RDTs compared to microscopy, indicate that RDTs are effective for detection of malaria infections for routine case management and surveillance of malaria in this area with confirmed ART-R; and can be utilized in the ongoing plans to develop a response to ART-R.

## Background

Malaria remains a major public health threat, especially among under-fives and pregnant women in Sub-Saharan Africa (SSA) including Tanzania [1]. In 2022, 93.6% of global malaria cases and 95.4% of the deaths due to malaria were reported in the World Health Organization’s (WHO) African region (WHO - Afro), where 78.1% of all malaria deaths in this region were among under-fives[2]. Tanzania is among the countries with the highest burden, it reported 4.4% of all global malaria deaths in 2022 [2]. Malaria in Tanzania is caused by three species of Plasmodium; *Plasmodium falciparum, Plasmodium ovale spp, and Plasmodium malariae,* with the majority of the infections (>85%) caused by *P. falciparum* [3–6]. Malaria infections due to *Plasmodium vivax* have been sporadically reported, but this parasite species is less prevalent due to the absence of Duffy antigen (among African populations), the binding site for *P. vivax [7]*. Despite the efforts that have been made to control malaria in Tanzania, challenges such as the emergence and spread of insecticide resistance in the vectors [8], antimalarials-resistant *P. falciparum [9]*, histidine-rich protein 2/3 (*hrp2/3*) gene deletions [10] and the emergence of invasive *Anopheles stephensi* vectors threaten the progress made in the past two decades[11].

For effective case management, WHO recommends parasitological confirmation of all suspected malaria cases by rapid diagnostic tests (RDTs) or microscopy before initiating treatment with artemisinin-based combination therapies (ACTs) [12]. Currently, RDTs are the primary diagnostic tool for malaria in Tanzania [13]. Their wide use is due to simplicity, short turn-around time, limited infrastructure requirements, and cost-effectiveness [14]. However, their performance is affected by various factors such as storage conditions, parasitemia, type of antigen, and operator skills [15]. Studies show that most RDTs tend to be less sensitive at low parasite densities (such as parasitaemia less than 100 asexual parasites/μL), thus missing low-density and chronic latent infections, especially in asymptomatic populations, particularly in low-transmission settings [16,17]. Different antigens are used in RDTs including lactate dehydrogenase (LDH), Aldolase, (Aldolase), and *P. falciparum* histidine-rich protein 2 (*pf*HRP2) [18]. Although *pf*HRP2-based RDTs are widely used due to their sensitivity, stability, and abundance, their accuracy may be limited by mutant or *hrp2/3* gene deletions [10], and or prozone effect [19,20]), thus leading to false negative results [21]

For more than a century, microscopy has remained the gold standard for malaria diagnosis due to its ability for detection and visualization, differentiate parasite species, detection parasite stages (sexual or asexual forms), and quantification of malaria parasites in blood smears [22]. While it offers high specificity and the ability to determine parasite species and quantify parasitemia, its sensitivity can be influenced by factors such as the quality of staining reagents and the skills of microscopists [23]. The limit of detection (LOD) of the expert microscopist can be as low as five (5) parasites/μL, while the average LOD for most microscopists ranges from 50 to 100 parasites/μL [24]. Other limitations of microscopy include the demand for high-quality microscopy which is well maintained, a laboratory facility, reliable electricity and reagents (fixing and staining reagents, and filtered water at the correct pH) [23,25]. Due to these limitations, most malaria-endemic countries deployed RDTs and have been using them for parasitological confirmation and supporting treatment using ACTs which significantly contributed to reducing the burden of malaria over the past two decades.

Malaria diagnosis by molecular techniques or nucleic acid detection methods is a highly sensitive and specific method that detects, amplifies, and quantifies DNA specific to malaria parasites [26]. Different methods have been developed including quantitative PCR (qPCR) and offer accurate and rapid detection of the parasites, even at low parasite densities (1-5 asexual parasites/μL), and can accurately differentiate different *Plasmodium* species, as well as identify mixed or complex infections [16,27,28]. These methods have high sensitivity and specificity which enable them to identify infections in different groups including asymptomatic reservoirs that are missed by conventional diagnostic methods (microscopy and RDTs), and can provide a more accurate assessment of malaria prevalence within a population [4,29]. Additionally, molecular techniques facilitate the detection and monitoring of antimalarial drug resistance by detecting genetic markers associated with resistance. They are also useful in efficacy studies of antimalarial drugs for differentiating recrudescent from new infections and thus establishing the efficacy of antimalarials [30]. However, nucleic acid detection methods have not been widely used in malaria-endemic countries including Tanzania, because they have some major limitations such as infrastructure requirements, lack of skilled experts, and high purchasing and operational costs [31].

In areas with high transmission, infections in the communities (mainly among asymptomatic individuals) play an important role in malaria transmission [32]. These individuals who remain in the community without health-care harbor parasites infections, more often with low levels of parasitemia that are difficult to detect by conventional diagnostic methods like RDTs and microscopy. This makes detection and targeting of these community infections difficult [21,33]. In addition, there is a paucity of data on how routine diagnostic methods such as RTDs and microscopy perform in the detection of infections among community members during routine surveillance of malaria. The problem is potentially higher in asymptomatic community members, who remain untreated but with high potential of sustaining transmission. In areas with biological threats such as artemisinin partial resistance (ART-R), it is critical to deploy and use highly sensitive tests as part of the response strategy to prevent the spread of resistant parasites. This study aimed to determine the performance of RDTs and microscopy using qPCR as a reference method for the detection of malaria parasites in community members (with or without symptoms) in Kyerwa district of Kagera region, an area where ART-R has been recently confirmed [9]. The findings of this study provide evidence for the potential use of RDTs and microscopy in the surveillance and targeting of community infections (mainly asymptomatic individuals) as part of the response to ART-R in Tanzania.

## Methods

### Study design and sites

The data and samples used in this study were obtained from a community cross-sectional survey (CSS) that was conducted in five villages in Kyerwa district of Kagera region as described earlier [34]. This was part of the main project on molecular surveillance of malaria in Mainland Tanzania (MSMT) which has been implemented in regions with varying endemicity since 2021 [3,4,34]. The five study villages (Kitoma, Kitwechenkura, Nyakabwera, Rubuga and Ruko) are located in Kyerwa district, which is among the eight councils of Kagera region as previously described [34]. The villages were selected based on recent research findings which showed that some areas of Kagera region have high levels of parasites with mutations associated with ART-R [9].

### Study population, participant enrollment, and data collection

In this study, community members aged 6 months and above were enrolled. The participants reside in the five study villages of Kyerwa district that are part of the longitudinal component of the MSMT project. The participants were asked and provided informed consent to participate in the study. Details of the procedures for enrollment of participants in the CSS were fully described in a recently published paper [34]. Briefly, demographic, anthropometric, clinical, and parasitological data were collected using a questionnaire developed and run-on tablets installed with an Open Data Kit (ODK) software version 4.2 [34]. Participants were appropriately identified using their permanent identification numbers (IDs), interviewed to collect demographic and anthropological data, clinically assessed and examined for any illness, and tested for malaria using RDTs as described earlier [34].

### Sample size and sample collection

The CSS aimed to recruit and obtain blood samples from selected individuals who voluntarily and were conveniently enrolled from a population of 17519 residing in 4144 households, as described earlier [34]. Through a convenient and non-random sampling which was used as described earlier [34], 4454 out of 17519 (25.4%) individuals residing in 768 households were enrolled. Enrolled participants provided finger prick blood for RDTs, thick and thin blood smears for detection of malaria parasites by microscopy, and dried blood spots (DBS) on filter papers (Whatman No. 3, GE Healthcare Life Sciences, PA, USA) for laboratory analyses. All samples were collected from participants using the standard operating procedures (SOPs) of the MSMT project. Briefly, each DBS had three spots, each with about 20 mm diameter equivalent to about 50μl of blood [3]. DBS samples were air-dried and packed in zipper bags with silica gel to prevent moisture and fungal infestation. Thick and thin blood smears were prepared in the field, air-dried, and thin films were fixed with absolute methanol. The slides were stained on the same or the following day with 5% Giemsa solution for 45 minutes, and packed in slide storage boxes [35]. All samples were stored at room temperature in the field before delivery to the Genomics Laboratory at the National Institute for Medical Research (NIMR), in Dar es Salaam, for subsequent processing and analyses.

### Malaria diagnosis using RDT

Detection of malaria infections was done using RDTs under field conditions in which Finger-prick blood was collected from all enrolled participants. Two brands of RDTs were used, Abbott Bioline Malaria Ag Pf/Pan (Abbott Diagnostics Korea Inc., Korea) and Smart Malaria Pf/Pan Ag Rapid Test (Zhejiang Orient Gene Biotech Co. Ltd, China). The RDTs used in the CSS had PfHRP-2 and pLDH antigens. The tests were performed and interpreted following the manufacturers’ instructions [36]. RDT results were sent to clinicians to make a final diagnosis in case malaria infections were suspected. Individuals with positive results but without a history of using antimalarial drugs in the last seven days were treated according to the national guidelines for the diagnosis and treatment of malaria [37], while other illnesses were appropriately treated as per national guidelines [38].

### Detection of malaria parasites by microscopy

The collected blood smears were read at the NIMR Genomics Laboratory after the completion of field activities. Two experts, WHO-certified microscopists read the blood smears for detection of malaria parasites, identification and quantitation of asexual and sexual (gametocytes) stages, and detection of different *Plasmodium* species as described earlier [23]. In case of discrepancy, a third reading was performed by an independent microscopist blinded to the results of the first two readers. In all positive smears, asexual and sexual parasites were counted against 200 and 500 white blood cells (WBCs), respectively. Parasite density was obtained by multiplying the parasite counts by 40 for asexual and 16 for sexual parasites, assuming each microliter of blood contained 8000 WBCs [39]. A blood slide was considered negative for Plasmodium species if no parasites were detected in at least 200 oil-immersion, high-power fields on the thick film. Quality control of smears was done as previously described [23].

### DNA extraction and detection of malaria parasites using real-time qPCR

DNA was extracted from three punches of DBS, and each punch had 6mm, representing approximately 25μl of blood. The extraction was done using 0.5% Chelex-Tween 20 (Bio-Rad Laboratories, Hercules, CA, USA), and the final DNA was eluted in a volume of approximately 100μl of nuclease-free water as previously described [3]. Species-specific qPCRs targeting 18S ribosomal Ribonucleic acid (rRNA) subunit were performed as described earlier [3], and the reactions were run on the Bio-Rad CFX 96 Opus real-time PCR detection system (Bio-Rad, California, USA), with CFX Maestro software version 2.2. A separate qPCR assay was run for each species: *P. falciparum, P. malariae,* and *P. ovale spp*. (simultaneously detecting both *P. ovale curtis* and *P. ovale wallikeri*) in a final volume of 12.5uL reaction mixture containing 10uL of master mix and 2.5uL of DNA template. qPCR was run for 40 cycles for all species except *P. malariae* which was run for 45 cycles, and quantification of the parasites was done by running standard curves using 10-fold serial dilution of engineered plasmids as earlier described [3]. Analysis of *P. vivax* was not done because it was rarely detected in our recent studies [3,4], but this will be done in the future using a pooling strategy that is being optimized.

### Data management and analysis

All clinical and parasitological data were collected using electronic data capture tools installed on tablets using ODK software version 4.2. The data were transmitted to the central server at NIMR in Dar es Salaam in real-time or once the internet connection was working, and they were validated daily concurrently with field activities. After completion of field activities, the data were downloaded into Microsoft Excel for further cleaning. Data analysis was done using STATA software version 13 (StataCorp, Texas, USA) and R software v4.4.1. Descriptive statistics including frequencies, percentages, medians, and interquartile ranges (IQRs) were used to summarize the data. Sensitivity, specificity, positive and negative predictive values, and accuracy of RDTs and microscopy were determined using 2×2 contingency tables, with qPCR as the gold standard [40]. For the three diagnostic techniques (RDTs, microscopy, and qPCR), Pearson Chi-squared test was used to assess the differences in malaria prevalence across the three age groups (under -fives, school children aged 5 - <15 years, and adult individuals with ≥15 years old), sex, history of fever in the past 48 hours, fever at presentation (axillary temperature ≥37.5°C), and area of residence [16,23]. The accuracy of RDTs and microscopy when compared to qPCR (defined as correct results with negative or positive tests divided by all results) were also determined as previously described [41]. Predictors of risk of obtaining false negative RDTs and microscopy results as determinants of sensitivities of RDTs and microscopy were computed using a multivariate logistic regression model, with adjustments for sex, age groups, history of fever in the past 48 hours, fever at presentation, parasite densities (<100, 100-1000, 1001-5000, 5001-10000, and >10000 asexual parasites/µL), and area of residence. For predictors of risk of obtaining false positive RDTs and microscopy results as determinants of the specificity of RDTs and microscopy, adjustments were done for sex, age groups, history of fever in the past 48 hours, fever at presentation, and area of residence. All results with a p-value <0.05 were considered significant. Concordance between the three methods was calculated using Cohen’s Kappa coefficient (κ), and *κ* < 0.20 indicated poor agreement, 0.21–0.40 was considered to be fair, 0.41–0.60 was moderate, 0.61–0.80 was good agreement, 0.81–0.99 was very good and 1.00 indicated perfect agreement [16].

## RESULTS

### Baseline characteristics of the population

The CSS which collected the data and samples used in this study was undertaken from 14^th^ July to 02^nd^ August 2023, and enrolled 4454 individuals from the five villages of Kyerwa district in Kagera region. The data and samples from all participants were available and were used in this study. The median age of study participants was 14 (IQR = 6.7 – 36.0) years; 59.3% (n = 2643/4454) were females, and the rest were males (40.7%, n = 1811/4454). Of all individuals, 48.2% (n = 2146/4454) were aged ≥15 years, while under-fives accounted for 18.7% (n = 835/4454) and 33.1% (n = 1473/4454) were school children (aged 5 - <15 years). Among the five villages, Nyakabwera had 27.9% (n = 1243/4454) of the study participants and Rubuga had 21.9% (n = 974/4454). Each of the three remaining villages had less than 20.0%, but had above 15.0% of the participants (Kitwechenkura with 17.3%, n = 769/4454; Ruko had 17.0%, n = 759/4454; and Kitoma had 15.9%, n = 709/4454). Overall, 30.1% (n = 1341/4454) of the participants had a history of fever in the past 48 hours before the survey, while only 3.1% (n =136/4454) had fever at presentation (with axillary temperature ≥37.5°C). Among all participants, 6.5% (n = 289/4454) had a history of medication use in the past seven days, and 89.3% (n = 258/289) of these reported that they used Artemether-Lumefantrine (AL) for the treatment of uncomplicated malaria (**Table 1**). Other antimalarial drugs reported to have been used included injectable artesunate (by 1.0%, n = 3/289), Metakelfin (by 0.7%, n = 2/289), Quinine (0.3%, n = 1/289), and other unspecified antimalarial drugs (0.3%, n = 1/289).

**Table 1.** Baseline characteristics of study participants.

| Characteristics | Kitoma | Kitwechenkura | Nyakabwera | Rubuga | Ruko | Total |
| --- | --- | --- | --- | --- | --- | --- |
| Examined, n (%) | 709 (15.9) | 769 (17.3) | 1243 (27.9) | 974 (21.9) | 759 (17.0) | 4454 |
| Sex, n (%) |  |  |  |  |  |  |
| Female | 393 (55.4) | 459 (59.7) | 749 (60.3) | 584 (60.0) | 458 (60.3) | 2643 (59.3) |
| Male | 316 (44.6) | 310 (40.3) | 494 (39.7) | 390 (40.0) | 301 (39.7) | 1811 (40.7) |
| Age in years, median (IQR) | 16 (8-38) | 15 (5-37) | 14 (6-35) | 12 (5-33) | 14(7-36) | 14.1 (6.7-36.0) |
| Age group (years), n (%) |  |  |  |  |  |  |
| <5 | 154 (21.7) | 131 (17.0) | 205 (16.5) | 180 (18.5) | 165 (21.7) | 835 (18.8) |
| 5 - <15 | 205 (28.9) | 230 (29.9) | 413 (33.2) | 351 (36.0) | 274 (36.1) | 1473 (33.1) |
| 15+ | 350 (49.4) | 408 (53.1) | 625 (50.3) | 443 (45.5) | 320 (42.2) | 2146 (48.2) |
| History of fever in the past 48 hours, n (%) |  |  |  |  |  |  |
| Yes | 296 (41.8) | 163 (21.2) | 130 (10.5) | 436 (44.8) | 316 (41.6) | 1341 (30.1) |
| No | 413 (58.3) | 606 (78.8) | 1113 (89.5) | 538 (55.2) | 443 (58.4) | 3113 (69.9) |
| Fever at presentation (temp ≥ 37.5°C) | 32 (4.5) | 21 (2.7) | 23 (1.9) | 41 (4.2) | 21 (2.8) | 138 (3.1) |
| History of use of any medication in the past seven days |  |  |  |  |  |  |
| Yes | 60 (8.5) | 54 (7.0) | 34 (2.7) | 64 (6.6) | 77 (10.1) | 289 (6.5) |
| No | 6549 (91.5) | 715 (93.0) | 1209 (97.3) | 910 (93.4) | 682 (89.9) | 4195 (94.5) |
| History of AL use in the past seven days |  |  |  |  |  |  |
| Yes | 55 (91.7) | 46 (85.2) | 29 (85.3) | 62 (96.9) | 66 (85.7) | 258 (89.3) |
| No | 5 (8.3) | 8 (14.8) | 5 (14.7) | 2 (3.1) | 11 (14.3) | 31 (10.7) |

**n: Number of observations; temp: Axillary temperature ( ≥ 37.5°C); IQR: Inter-quartile range; AL: Artemether-Lumefantrine**

### Prevalence of malaria by RDTs, microscopy, and qPCR

All enrolled participants were tested with the three methods, and 44.4% (n=1979/4454) had positive results by RDTs, while the prevalence was 32.1% (n = 1431/4454) by microscopy and 39.8% (n=1771/4454) by qPCR. The differences in the prevalence by the three methods were statistically significant (p<0.001). The highest prevalence of malaria infections by RDT (68.5%, n=520/759) and microscopy (51.6%, n=392/759) was in Ruko, while by qPCR, the highest prevalence was in Rubuga (55.9%, n=544/974); with significantly high variations among the villages (p<0.001). Nyakabwera had the lowest prevalence by all methods; RDTs (14.5%, n= 180/1243), microscopy (9.3%, n=116/1243) and qPCR (13.9%, n=173/1243); and the differences were statistically significant (p<0.001 for all tests) (**Table 2**).

**Table 2.** Prevalence of malaria parasites by RDT, microscopy and qPCR.

| Variable | # Positive by RDT, n (%) | # Positive by microscopy, n (%) | # Positive by qPCR, n (%) |
| --- | --- | --- | --- |
| Overall (n=4454) | 1979 (44.4) | 1431(32.1) | 1771(39.8) |
| Sex |  |  |  |
| Female | 1089 (41.2) | 808 (30.6) | 992 (37.5) |
| Male | 890 (49.1) | 623 (34.4) | 779 (43.0) |
| p-value | <0.001 | 0.007 | <0.001 |
| Age group (years) |  |  |  |
| <5 | 452 (54.1) | 259 (31.0) | 335 (40.1) |
| 5 - <15 | 874 (59.3) | 681 (46.2) | 794 (53.9) |
| ≥15 | 653 (30.4) | 491 (22.9) | 642 (29.9) |
| p-value | <0.001 | <0.001 | <0.001 |
| Village |  |  |  |
| Kitoma | 440 (62.1) | 345 (48.7) | 395 (55.7) |
| Kitwechenkura | 241 (31.3) | 176 (22.9) | 259 (33.7) |
| Nyakabwera | 180 (14.5) | 116 (9.3) | 173 (13.9) |
| Rubuga | 598 (61.4) | 401 (41.2) | 544 (55.9) |
| Ruko | 520 (68.5) | 392 (51.7) | 400 (52.7) |
| p-value | <0.001 | <0.001 | <0.001 |
| History of fever past 48 hours |  |  |  |
| Yes | 1186 (88.4) | 846 (63.1) | 1021 (76.1) |
| No | 793 (25.5) | 584 (18.8) | 750 (24.1) |
| p-value | <0.001 | <0.001 | <0.001 |
| Fever at presentation (axillary temp ≥ 37.5°C) |  |  |  |
| Yes | 108 (79.4) | 93 (68.4) | 101 (74.3) |
| No | 1871 (43.3) | 1338 (31.0) | 1670 (38.7) |
| p-value | <0.001 | <0.001 | <0.001 |
| History of using AL in the past seven days |  |  |  |
| Yes | 249 (96.5) | 97 (37.6) | 165 (64.0) |
| No | 1730 (41.2) | 1333 (31.8) | 1606 (38.3) |
| p-value | <0.001 | 0.053 | <0.001 |

**n: Number of observations; RDTs: Rapid diagnostic tests; qPCR: Quantitative Polymerase Chain Reaction; AL: Artemether-Lumefantrine; temp: Axillary temperature; #: number of individuals with positive results**

Using all three test methods, males and school children had significantly higher prevalence (p<0.05 for all comparisons, except with RDT for under-fives vs. school children) (Table 2; Fig. 2A and Fig. 2B). Among individuals with a history of fever within the past 48 hours, the majority tested positive by all methods (88.4%, n = 1186/1341 by RDTs; 63.2%, n = 847/1341 by microscopy; and 76.1%, n = 1021/1341 by qPCR). The prevalence was also higher for those with fever at presentation (axillary temperature ≥ 37.5°C), with 79.4% (n = 108/136) by RDTs, 68.4% (n = 93/136) by microscopy, and 74.3% (n = 101/136) by qPCR. Of the participants who reported that they used AL within the past seven days, 96.5% (n = 249/258) had positive results by RDTs; 37.6% (n = 97/258) were positive by microscopy while 64.0% (n = 165/258) had positive results by qPCR (**Table 2**).

**Figure 1:**
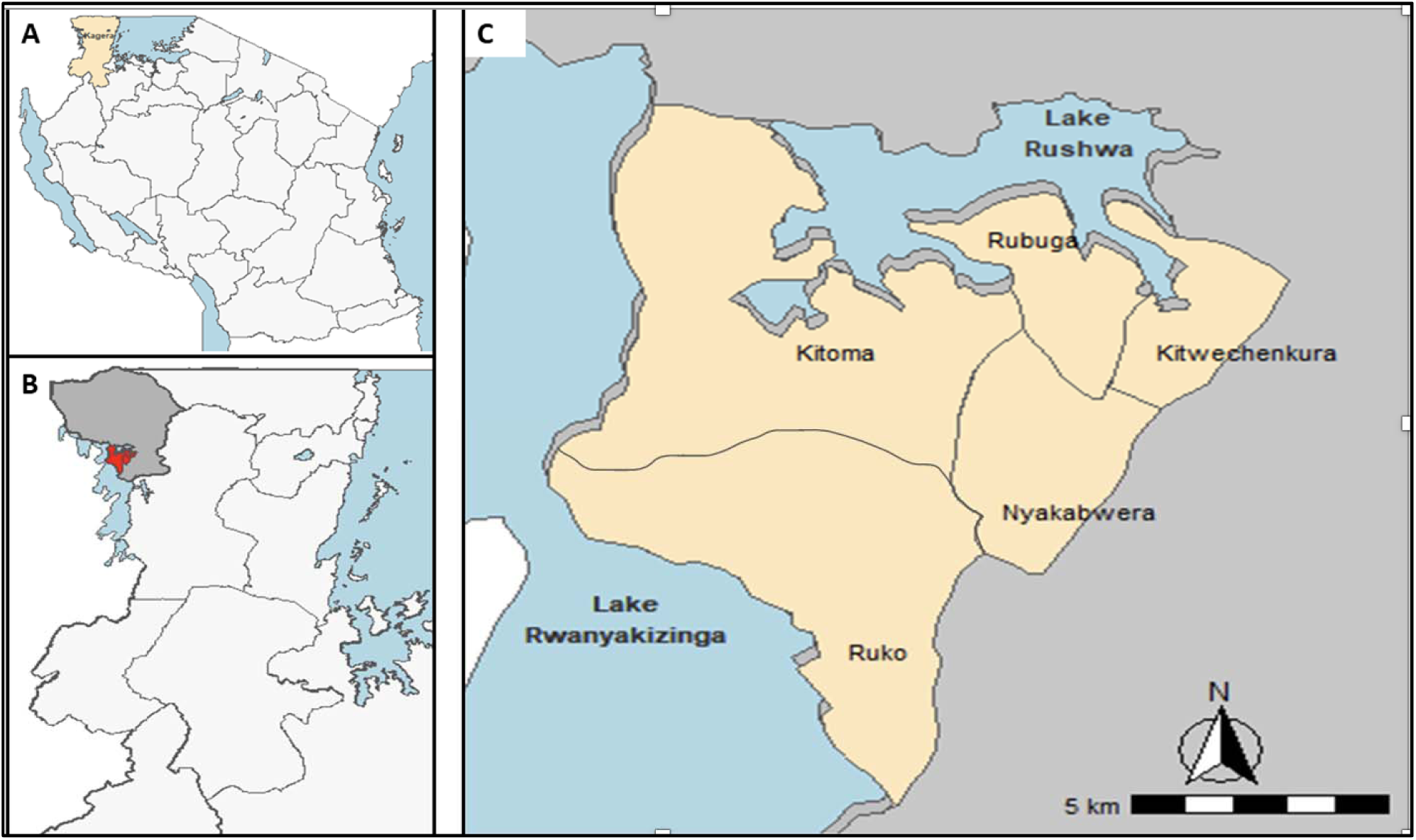
(A) Map of Tanzania showing the 26 regions including Kagera (gold), (B) study area in Kyerwa district (red), and (C) study villages (gold).

**Figure 2A.**
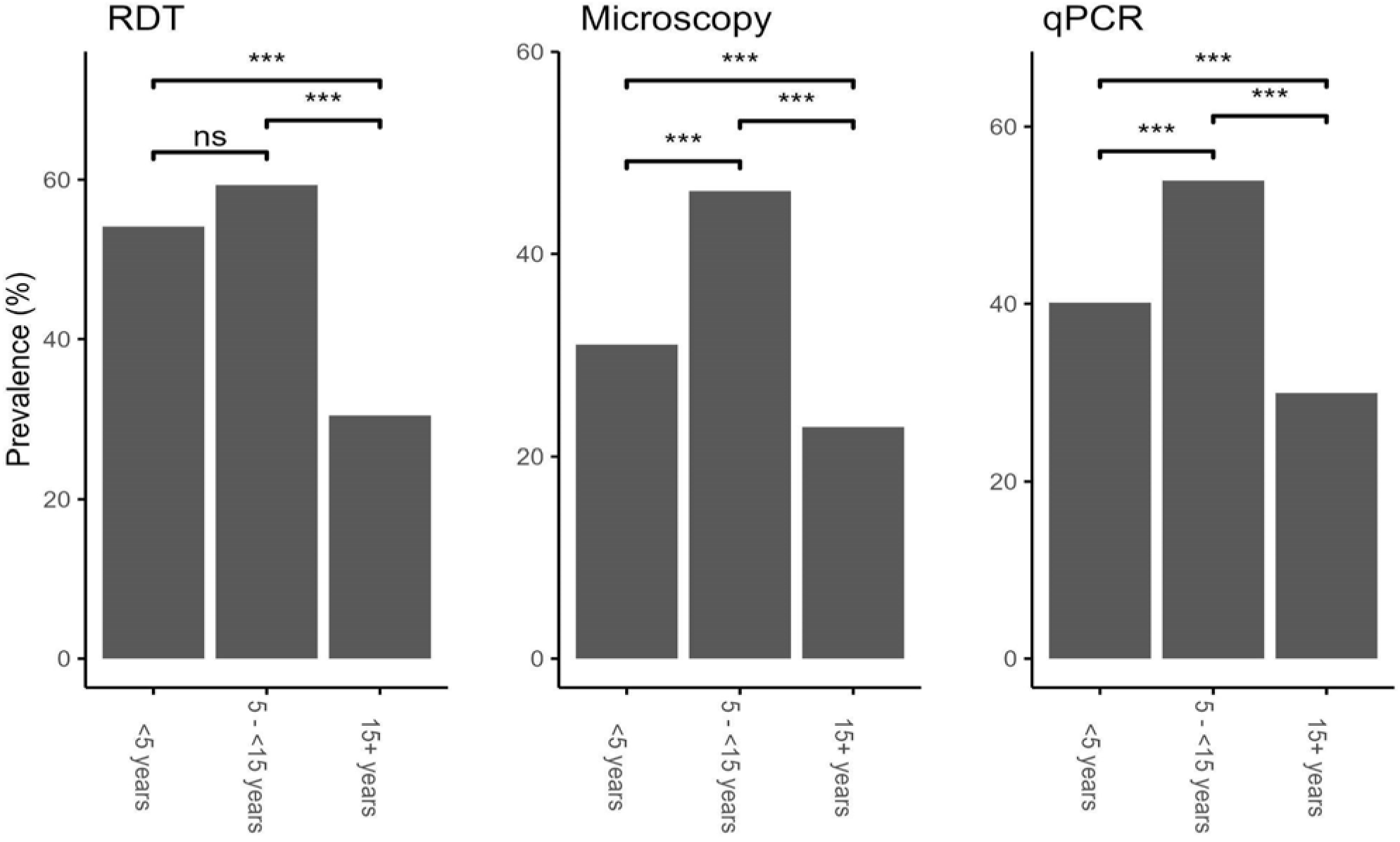
Prevalence of malaria infections by RDTs, microscopy, and qPCR among individuals of different age groups. *** p=0.001, **p<0.01, ns= Not significant

**Figure 2B.**
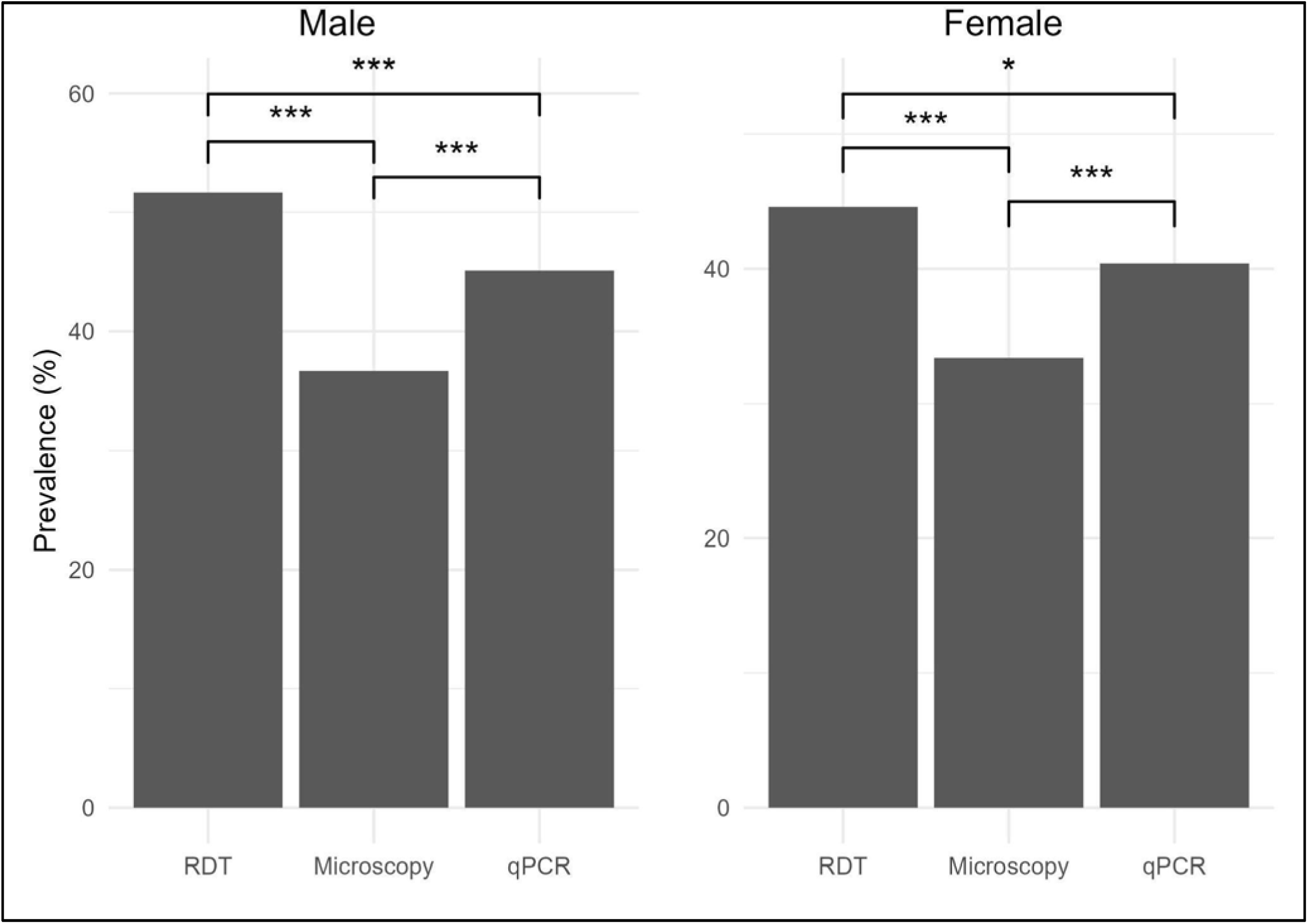
Prevalence of malaria infections by RDTs, microscopy, and qPCR among male and female participants. *** p=0.001, **p<0.01, *p<0.05

### Prevalence of *Plasmodium falciparum*, non-falciparum species and parasitaemia

Based on microscopy, the prevalence of *P. falciparum, P. malariae,* and *P. ovale* mono-infections was 28.7% (n = 1279/4454), 0.2% (n = 7/4454), and 0.3% (n = 12/4454), respectively. Mixed infections occurred in 3.0% (n = 133/4454) of the samples, and these included double infections, with 2.1% (n = 93/4454) of *P. falciparum/P. malariae* and 0.9% (n = 39/4454) with *P. falciparum/P. ovale*, and triple infections of *P. falciparum/P. malariae/P. ovale* which occurred in one sample (0.02%; n = 1/4454). By qPCR, the prevalence of *P. falciparum*, *P. malariae,* and *P. ovale* mono-infections was 35.3% (n = 1572/4454), 0.4% (n = 16/4454), and 0.5% (n = 24/4454), respectively. The proportion of samples with positive results due to mixed infections of different *Plasmodium* species by qPCR was 3.6% (n = 159/4454). These mixed infections included double infections of *P. falciparum/P. malariae* at 0.9% (n = 40/4454)*, P. falciparum/P. ovale at 2.6*% (n = 114/4454), and *0.04% (n = 2/4454) had P. ovale/P. malariae,* while triple infections of *P. falciparum/P. malariae/P. ovale* occurred in 0.07% (n = 3/4454) of the samples (**Table 3**).

**Table 3.** Prevalence of Plasmodium species by microscopy and qPCR.

| Plasmodium species | microscopy, n (%) | qPCR, n (%) |
| --- | --- | --- |
| All species (n=4454) | 1431(32.1%) | 1771(39.8%) |
| *P. falciparum* | 1279 (28.7) | 1572 (35.3) |
| *P. ovale* | 12 (0.3) | 24 (0.5) |
| *P. malariae* | 7 (0.2) | 16 (0.4) |
| *P. falciparum*/*P. malariae* | 93 (2.1) | 40 (0.9) |
| *P. falciparum*/*P. ovale* | 39 (0.9) | 114 (2.6) |
| *P. ovale*/*P. malariae* | 0 (0) | 2 (0.04) |
| *P. falciparum*/*P. malariae*/*P. ovale* | 1 (0.02) | 3 (0.07) |

**n: Number of observations; qPCR: Quantitative Polymerase Chain Reaction**

The geometric mean parasite densities (GMPDs) by microscopy were 642 parasites/µL (95% CI = 570 - 723) for *P. falciparum,* 124 parasites/µL (95% CI = 97 - 160) for *P. malariae*, and 126 parasites/µL (95% CI = 82-194) for *P.ovale* spp (Fig. 3A). The parasite densities of *P. falciparum* were significantly lower (p<0.001) among adults aged ≥15 years (with the mean parasite density of 246 parasites/µL, 95% CI = 206-293), while the highest densities were among under-fives (with 1915 parasites/µL; 95% CI = 1398 - 2623), 209 parasites/µL (70 - 629), and 288 parasites/µL (165 - 504) for *P. falciparum*, *P. ovale,* and *P. malariae,* respectively. The GMPDs of *P. falciparum* were higher in males compared to females (with 759 parasites/µL, 95% CI = 635 - 908 for males *versus* 563 parasites/µL, 95% CI=481-660 for females, p=0.014) and in Ruko (847 parasites/µL, 95% CI = 679 - 1057, p=0.034) compared to other villages (Table 4). By qPCR, the GMPDs of *P. falciparum, P. ovale,* and *P. malariae* were 1180 parasites/µl (95% CI = 1032 - 1349), 44 parasites/µl (95% CI = 32 - 61), and 50 parasites/µl (95%CI = 29 - 89) (Fig 3B). The parasite densities of *P. falciparum* were lower among adults aged ≥15 years, with a mean parasite density of 566 parasites/µL, 95% CI = 460-697, p<0.001), compared to under-fives who had higher densities (with a mean parasite density of 2076 parasites/µL, 95% CI = 1462 - 2948, p<0.001). The mean parasite densities for *P. ovale* were also significantly higher among under-fives (with 118 parasites/µL, 95% CI = 59-236, p=0.002), while the GMPD of *P. malariae* in under-fives (67 parasites/µL, 95% CI=15-300, p=0.272) was also higher, but the differences among age groups were not statistically significant. Parasitemia due to *P. falciparum* was higher in males (with 1401 parasites/µL, 95% CI = 1143-1717, p=0.015) while with *P. malarie* and *P. ovale,* the differences in GMPD among females and males were not statistically significant (p=0.528, and p=0.225 for *P. malarie*, and *P. ovale*, respectively) (**Table 4**).

**Figure 3:**
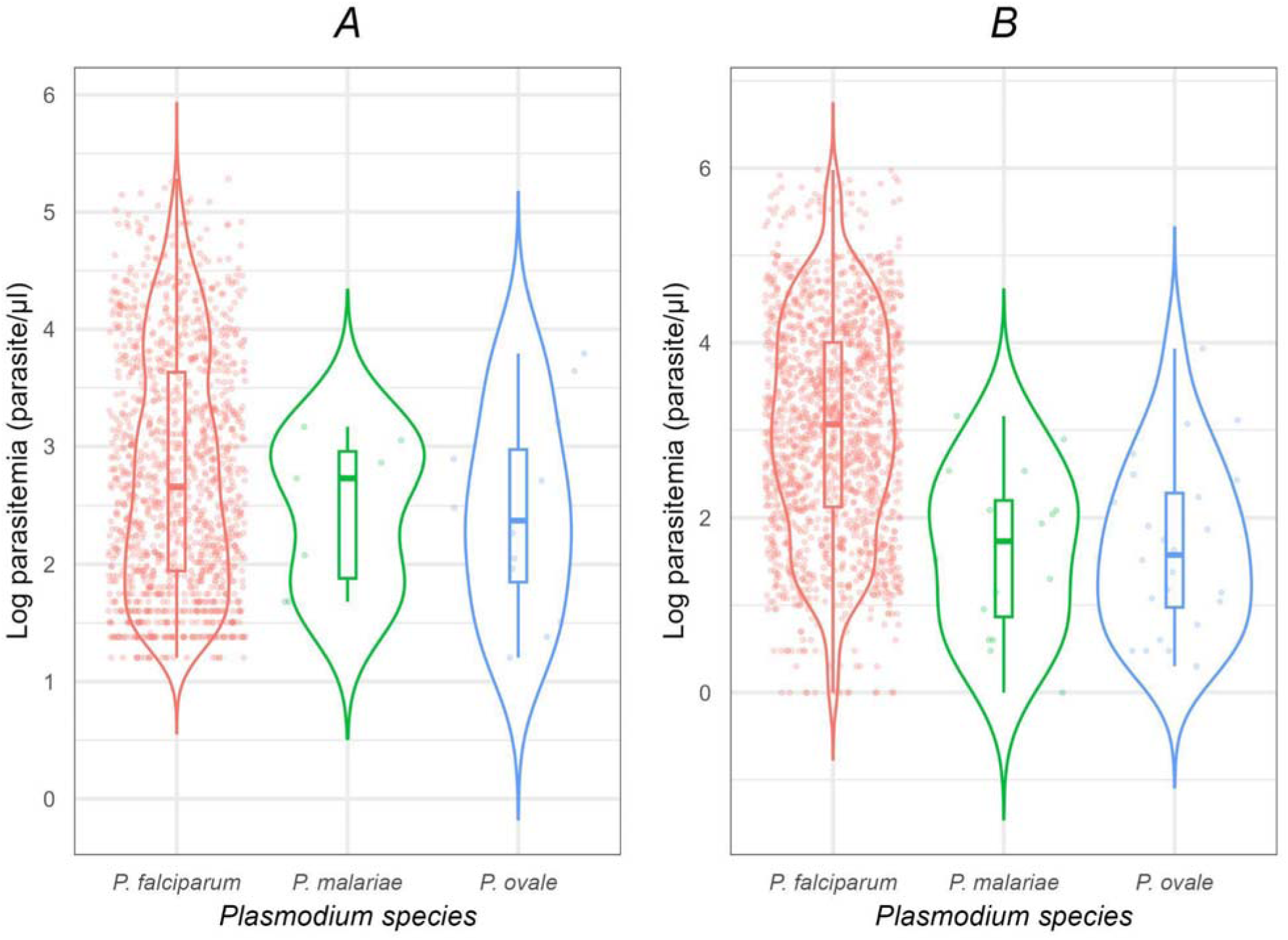
Violin plots representing the distribution of parasite densities detected for *P. falciparum, P. malariae, and P. ovale*. Panel A shows densities detected using microscopy, while Panel B presents those detected using PCR. The plots highlight the variability in parasite densities for each species, with the shape and width of the violins indicating the spread and frequency of the data points.

**Table 4.**
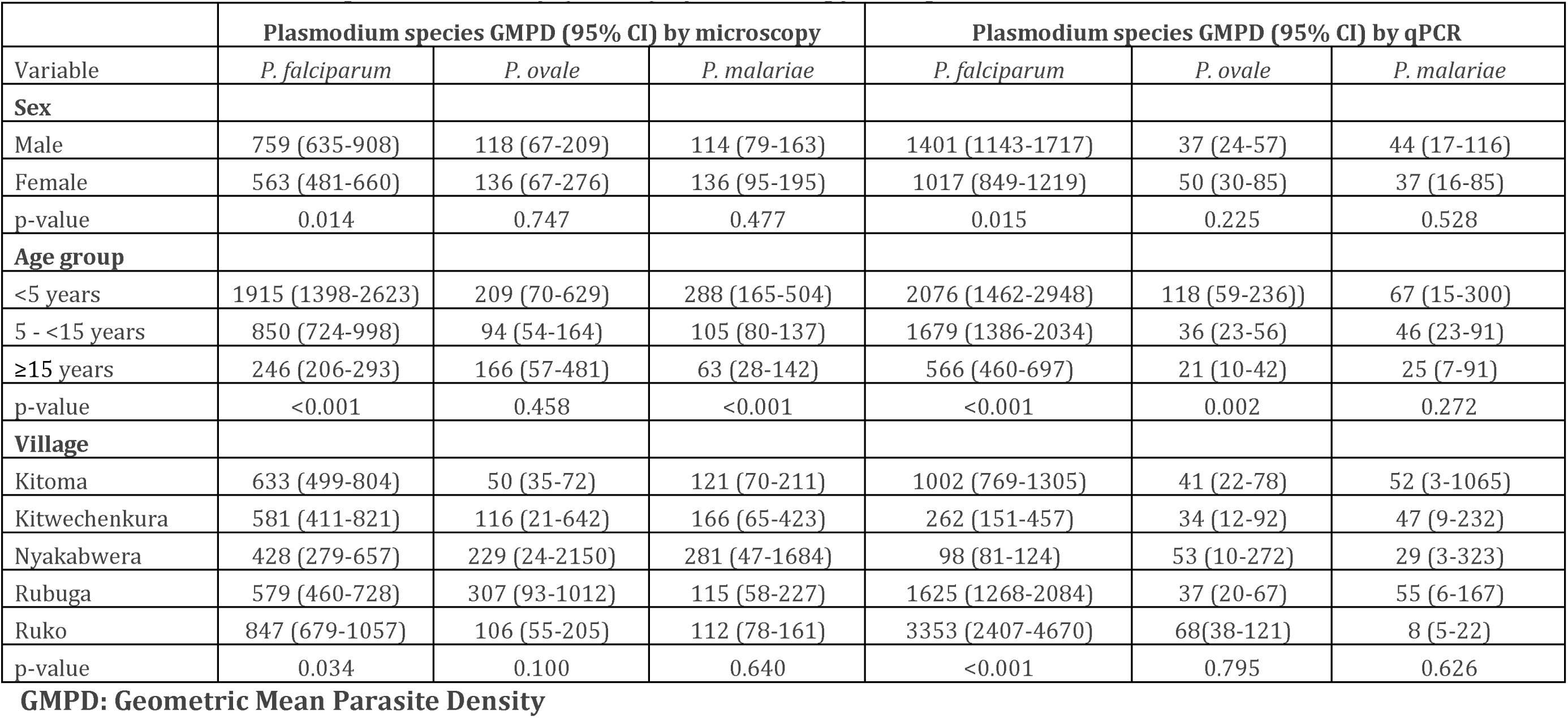
Geometric mean parasite density (GMPD) by microscopy and qPCR.

|  | **Plasmodium species GMPD (95% CI) by microscopy** |  |  | **Plasmodium species GMPD (95% CI) by qPCR** |  |  |
| --- | --- | --- | --- | --- | --- | --- |
| Variable | *P. falciparum* | *P. ovale* | *P. malariae* | *P. falciparum* | *P. ovale* | *P. malariae* |
| **Sex** |  |  |  |  |  |  |
| Male | 759 (635-908) | 118 (67-209) | 114 (79-163) | 1401 (1143-1717) | 37 (24-57) | 44 (17-116) |
| Female | 563 (481-660) | 136 (67-276) | 136 (95-195) | 1017 (849-1219) | 50 (30-85) | 37 (16-85) |
| p-value | 0.014 | 0.747 | 0.477 | 0.015 | 0.225 | 0.528 |
| **Age group** |  |  |  |  |  |  |
| <5 years | 1915 (1398-2623) | 209 (70-629) | 288 (165-504) | 2076 (1462-2948) | 118 (59-236)) | 67 (15-300) |
| 5 - <15 years | 850 (724-998) | 94 (54-164) | 105 (80-137) | 1679 (1386-2034) | 36 (23-56) | 46 (23-91) |
| ≥15 years | 246 (206-293) | 166 (57-481) | 63 (28-142) | 566 (460-697) | 21 (10-42) | 25 (7-91) |
| p-value | <0.001 | 0.458 | <0.001 | <0.001 | 0.002 | 0.272 |
| **Village** |  |  |  |  |  |  |
| Kitoma | 633 (499-804) | 50 (35-72) | 121 (70-211) | 1002 (769-1305) | 41 (22-78) | 52 (3-1065) |
| Kitwechenkura | 581 (411-821) | 116 (21-642) | 166 (65-423) | 262 (151-457) | 34 (12-92) | 47 (9-232) |
| Nyakabwera | 428 (279-657) | 229 (24-2150) | 281 (47-1684) | 98 (81-124) | 53 (10-272) | 29 (3-323) |
| Rubuga | 579 (460-728) | 307 (93-1012) | 115 (58-227) | 1625 (1268-2084) | 37 (20-67) | 55 (6-167) |
| Ruko | 847 (679-1057) | 106 (55-205) | 112 (78-161) | 3353 (2407-4670) | 68(38-121) | 8 (5-22) |
| p-value | 0.034 | 0.100 | 0.640 | <0.001 | 0.795 | 0.626 |

**GMPD: Geometric Mean Parasite Density**

### Performance of RDTs, microscopy, and qPCR for malaria detection

Of all participants (n = 4454), 1651 (37.1%, 95% CI = 35.3-38.9) were positive by both RDTs and qPCR, while 1328 participants (29.8%, 95% CI = 28.2-31.5) tested positive by both microscopy and qPCR (Table 5 and Fig. 4). With qPCR as the reference method, the sensitivity of RDTs and microscopy was 93.2% (95% CI = 92.0 - 94.4) and 75.0% (95% CI = 72.9 - 77.0), respectively. The specificity of RDTs was 87.8% (95% CI = 86.5 - 89.0), while that of microscopy was 96.2% (95% CI = 95.4 - 96.9) (Table 5 and Fig 4). The diagnostic accuracy of RDTs was 89.9% (95% CI = 89.0-90.8), and the accuracy was 87.7% (86.7-88.7) for microscopy. Microscopy showed a higher positive predictive value of 92.8% (95% CI = 91.4 - 94.0) compared to RDTs which had a positive predictive value of 83.4% (95% CI = 82.0 - 84.8). Conversely, the negative predictive value was higher for RDT than for microscopy, 95.2% (95% CI = 94.3 - 95.9) and 85.30% (95% CI = 84.3 - 86.3), respectively. Both RDTs and microscopy had better agreement with qPCR, with Cohen’s kappa values of 0.79 (95% CI = 0.78-0.81) and 0.74 (95% CI = 0.71-0.76), respectively (**Table 5**).

**Figure 4.**
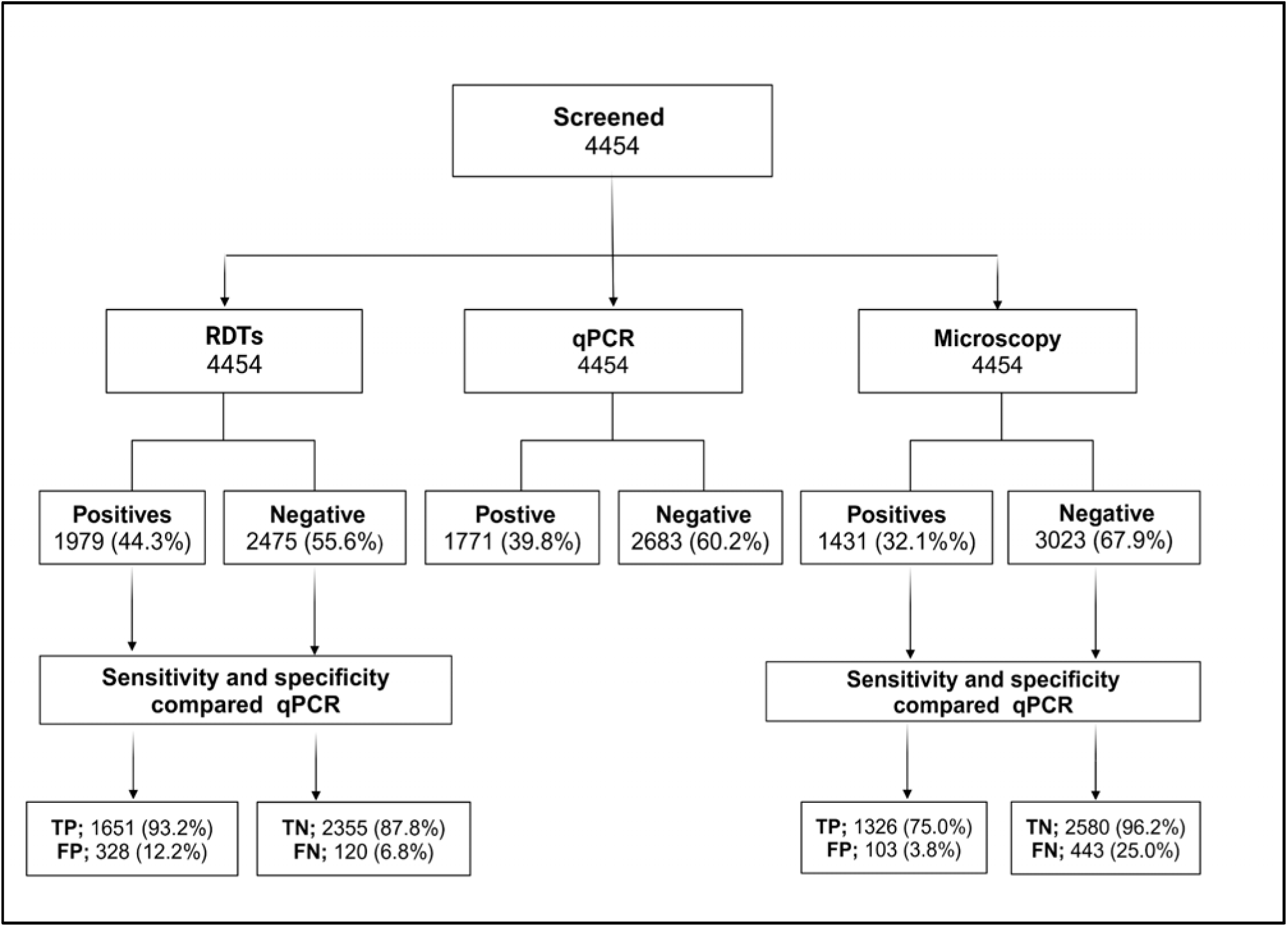
Flow chart showing tests performed and their diagnostic performances. TP: True Positive; FP: False Positive; TN: True Negative; FN: False Negative

**Table 5.**
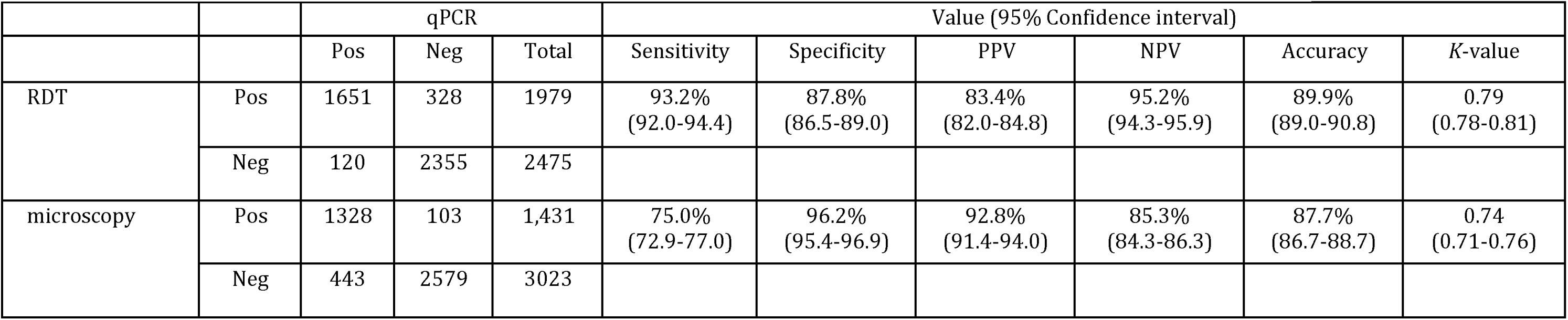
Performance of RDT and microscopy with qPCR as a reference method.

The sensitivities of RDTs and microscopy varied at different levels of parasitemia, with the lowest sensitivity of microscopy of 40.4% at <100 parasites/μL while for RDTs, the sensitivity at parasitaemia <100 parasites/μL was 76.6% (Fig 5). The sensitivity of both RDTs and microscopy increased with an increase in parasitemia, from 70.7% and 94.0% at 100-1000 parasites/μL to 92.3% and 100% at >5000-10000 parasites/μL by microscopy and RDTs, respectively. The sensitivity was over 99.6% at very high parasite densities (>10000 parasites/μL) (Fig 5). The sensitivity of both methods (RDTs and microscopy) was significantly higher at high parasitaemia (aOR >100, p<0.001) and among individuals with a history of fever (aOR > 1.30, p<0.001) (**Table 6A**). The specificity of RDTs was significantly affected by age (aOR >0.40, p<0.001), history of fever (aOR =0.08, p<0.001), and fever at presentation (aOR = 0.59, p<0.05) while the specificity of microscopy was not affected by any of the demographic and clinical variables (p>0.05) (**Table 6B**).

**Figure 5.**
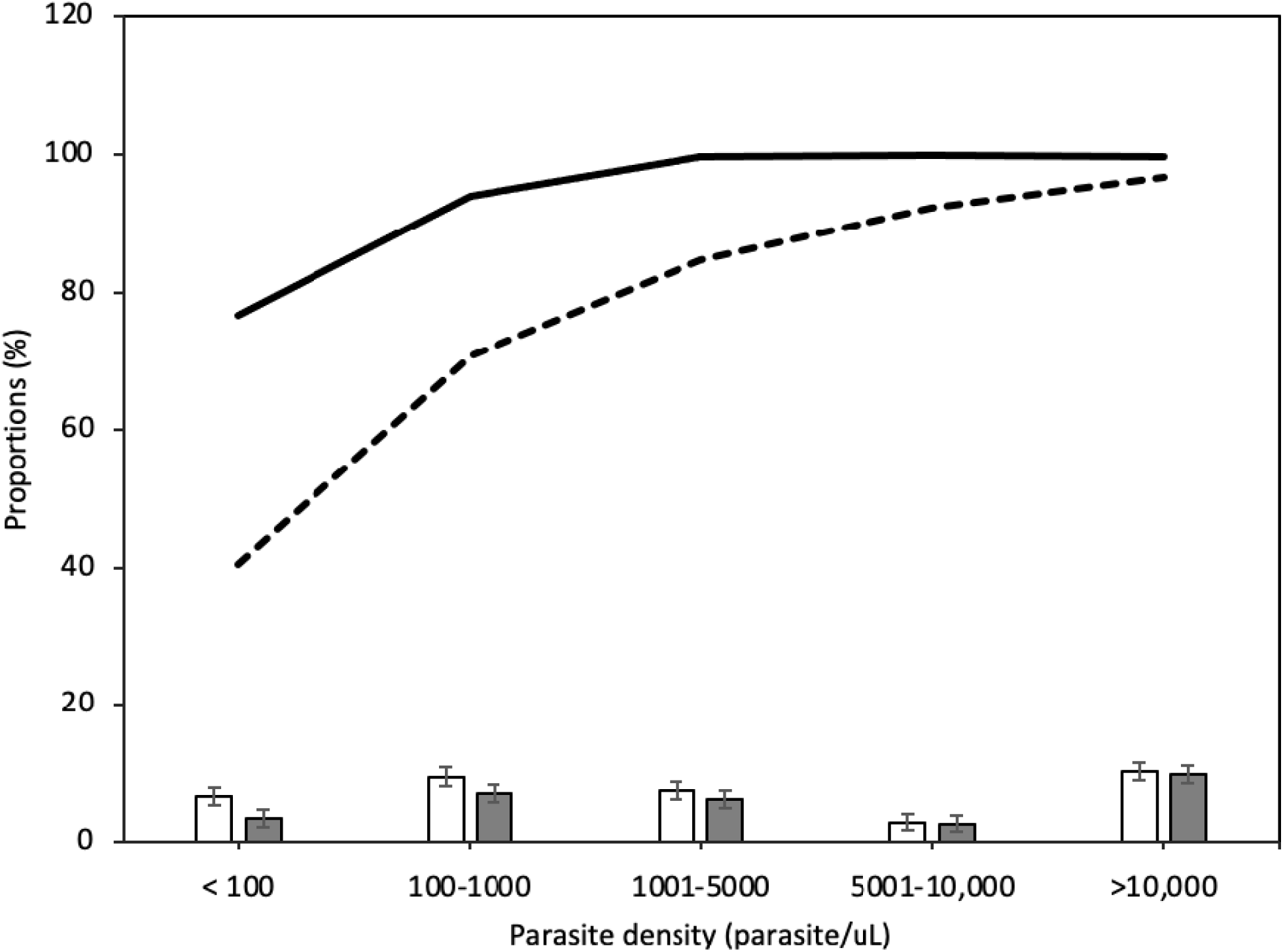
Sensitivity of RDTs and microscopy at different levels of parasitemia. Bars represent proportion of cases with positive results by different categories of parasite density, asexual parasites/μL (black bars =microscopy, n = 4454; and clear bars =RDTs, n = 4454); Solid line = sensitivity of RDTs and dotted line = sensitivity of microscopy.

**Table 6A.** Predictors of a risk sensitivity of RDT and microscopy results among individuals with positive results by qPCR.

|  | RDTs |  |  | Microscopy |  |  |
| --- | --- | --- | --- | --- | --- | --- |
| Variable/covariate | Sensitivity<br>(%, 95% CI) | Unadjusted OR<br>(95% CI) | Adjusted OR<br>(95% CI) | Sensitivity<br>(%, 95% CI) | Unadjusted OR<br>(95% CI) | Adjusted OR<br>(95% CI) |
| **Sex** |  |  |  |  |  |  |
| Male | 745/779 (95.6, 94.0-96.9) | Ref | Ref | 588/779 (75.5, 72.3-78.4) | Ref | Ref |
| Female | 906/992 (91.3, 89.4-92.9) | 0.48 (0.32-0.72) *** | 0.52 (0.31-0.86) * | 740/992 (74.6, 71.8-77.2) | 0.95 (0.77-1.18) | 1.14(0.86-1.51) |
| **Age group** |  |  |  |  |  |  |
| <5 years | 329/335(98.2, 96.1-99.2) | Ref | Ref | 245/335(73.1, 68.1-77.6) | Ref | Ref |
| 5 - <15 years | 768/794(96.7, 95.2-97.8) | 0.54 (0.20-1.24) | 0.53 (0.18-1.34) | 651/794 (82.0, 79.2-84.5) | 1.67(1.23-2.26) *** | 2.37<br>(1.58-3.55) *** |
| ≥15 years | 554/642 (86.3, 83.4-88.7) | 0.11 (0.04-0.24) *** | 0.18 (0.1-0.42)<br>*** | 432/642 (67.3, 63.6-70.8) | 0.76 (0.56-1.01) | 1.45 (0.98-2.15) |
| **History of fever** |  |  |  |  |  |  |
| No | 642/750 (85.6, 82.9-87.9) | Ref | Ref | 507/750 (67.6, 64.2-70.9) | Ref | Ref |
| Yes | 1009/1021 (98.8, 98.0-99.3) | 14.14<br>(8.05-27.27) *** | 8.78<br>(4.73-17.69) *** | 821/1021 (80.4, 77.9-82.7) | 1.97 (1.58-2.45) *** | 1.35 (1.02-1.79) * |
| **Fever at presentation** |  |  |  |  |  |  |
| No | 1550/1670(92.8, 91.5-94.0) | Ref | Ref | 1236/1670 (74.0, 71.976.1) | Ref | Ref |
| Yes | 101/101 (100, 96.3-100) | 1.29 (1.03-1.62) * | 1.03 (0.76-1.39) | 92/101 (91.1, 83.9-95.2) | 1.24 (1.09-1.42) ** | 1.05 (0.88-1.26) |
| **Parasite density** |  |  |  |  |  |  |
| <100 | 302/391 (77.2, 72.8-81.1) | Ref | Ref | 158/391 (40.4, 35.7-45.3) | Ref | Ref |
| 100-1000 | 426/454 (93.8, 91.2-95.7) | 1.39<br>(0.85-2.24) | 1.75<br>(0.98-3.14) | 321/454 (70.7, 66.4-74.7) | 2.45(1.60-3.79) *** | 2.81<br>(1.74-4.58) *** |
| 1001-5000 | 338/339 (99.7, 98.3-99.9) | 4.00 (2.34-6.99) *** | 5.36<br>(2.82-10.48) *** | 287/339 (84.7, 80.4-88.1) | 5.11(3.37-7.85) *** | 5.86<br>(3.66-9.52) *** |
| 5001-10000 | 129/129 (100, 97.1-100) | 44.21<br>(18.76-129.96) *** | 56.76<br>(21.76-180.9) *** | 119/129 (92.2, 86.3-95.7) | 14.11<br>(9.45-21.38) *** | 20.68<br>(13.0-33.61) *** |
| >10000 | 456/458 (99.6, 98.4-99.9) | 81.51<br>(24.63-504.50) *** | 104.67<br>(27.83-6927) *** | 443/458 (96.7, 94.7-98.0) | 70.92<br>(39.71-134.23) *** | 146.1<br>(76.5-294.4) *** |
| **Village of residence** |  |  |  |  |  |  |
| Kitwechenkura | 220/259(84.9, 80.1-88.8) | Ref | Ref | 167/259 (64.5, 58.5-70.1) | Ref | Ref |
| Nyakabwera | 154/173(89.0, 83.5-92.9) | 1.44 (0.81-2.63) | 6.30<br>(2.90-14.2) *** | 92/173 (53.2, 45.8-60.5) | 0.63 (0.42-0.93) * | 1.95 (1.16-3.28) * |
| Rubuga | 361/395(91.4, 88.2-93.7) | 1.88 (1.15-3.08) * | 4.17<br>(2.12-8.41) *** | 336/395 (85.1, 81.2-88.2) | 3.14 (2.16-4.59) *** | 9.27<br>(5.77-15.08) *** |
| Kitoma | 529/544(97.2, 95.5-98.3) | 6.25<br>(3.45-11.92) *** | 9.49<br>(4.47-21.2) *** | 374/544 (68.8, 64.7-72.5) | 1.21 (0.89-1.65) | 1.65 (1.11-2.46) * |
| Ruko | 387/400(96.8, 94.5-98.1) | 5.28<br>(2.83-10.47) *** | 14.6<br>(6.55-34.6) *** | 359/400 (89.8, 86.4-92.4) | 4.82 (3.22-7.34) *** | 14.8<br>(8.98-24.92) *** |

OR: Odds ratio, CI: Confidence interval, \*\*\*p<0.001, \*\*p<0.01, \*p<0.05

**Table 6B.**
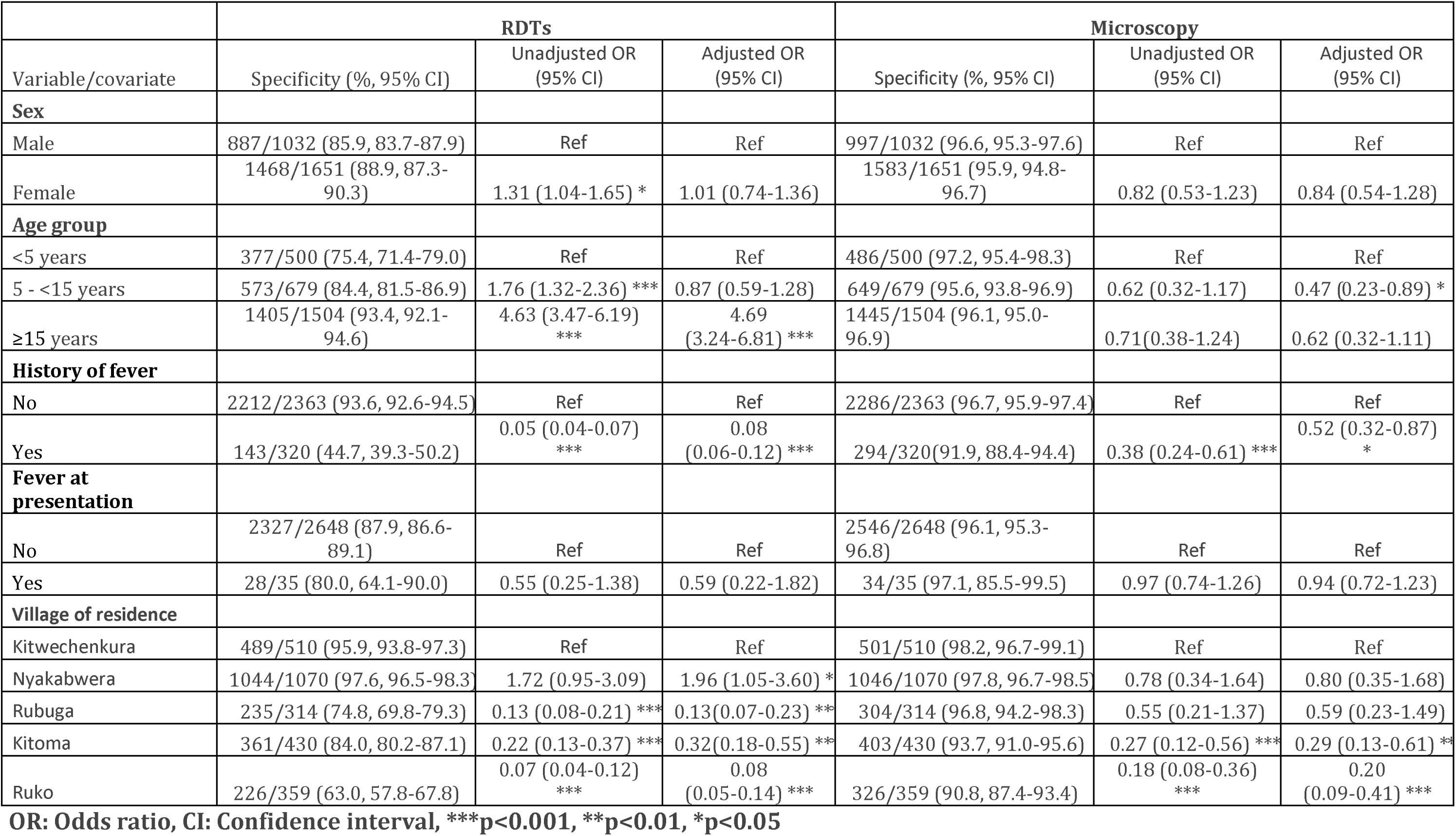
Predictors of specificity of RDT and microscopy results among individuals with negative results by qPCR.

|  | RDTs |  |  | Microscopy |  |  |
| --- | --- | --- | --- | --- | --- | --- |
| Variable/covariate | Specificity (%; 95% CI) | Unadjusted OR (95% CI) | Adjusted OR (95% CI) | Specificity (%; 95% CI) | Unadjusted OR (95% CI) | Adjusted OR (95% CI) |
| **Sex** |  |  |  |  |  |  |
| Male | 887/1032 (85.9, 83.7-87.9) | Ref | Ref | 997/1032 (96.6, 95.3-97.6) | Ref | Ref |
| Female | 1468/1651 (88.9, 87.3-90.3) | 1.31 (1.04-1.65) * | 1.01 (0.74-1.36) | 1583/1651 (95.9, 94.8-96.7) | 0.82 (0.53-1.23) | 0.84 (0.54-1.28) |
| **Age group** |  |  |  |  |  |  |
| <5 years | 377/500 (75.4, 71.4-79.0) | Ref | Ref | 486/500 (97.2, 95.4-98.3) | Ref | Ref |
| 5 - <15 years | 573/679 (84.4, 81.5-86.9) | 1.76 (1.32-2.36) *** | 0.87 (0.59-1.28) | 649/679 (95.6, 93.8-96.9) | 0.62 (0.32-1.17) | 0.47 (0.23-0.89) * |
| ≥15 years | 1405/1504 (93.4, 92.1-94.6) | 4.63 (3.47-6.19) *** | 4.69 (3.24-6.81) *** | 1445/1504 (96.1, 95.0-96.9) | 0.71(0.38-1.24) | 0.62 (0.32-1.11) |
| **History of fever** |  |  |  |  |  |  |
| No | 2212/2363 (93.6, 92.6-94.5) | Ref | Ref | 2286/2363 (96.7, 95.9-97.4) | Ref | Ref |
| Yes | 143/320 (44.7, 39.3-50.2) | 0.05 (0.04-0.07) *** | 0.08 (0.06-0.12) *** | 294/320(91.9, 88.4-94.4) | 0.38 (0.24-0.61) *** | 0.52 (0.32-0.87) * |
| **Fever at presentation** |  |  |  |  |  |  |
| No | 2327/2648 (87.9, 86.6-89.1) | Ref | Ref | 2546/2648 (96.1, 95.3-96.8) | Ref | Ref |
| Yes | 28/35 (80.0, 64.1-90.0) | 0.55 (0.25-1.38) | 0.59 (0.22-1.82) | 34/35 (97.1, 85.5-99.5) | 0.97 (0.74-1.26) | 0.94 (0.72-1.23) |
| **Village of residence** |  |  |  |  |  |  |
| Kitwechenkura | 489/510 (95.9, 93.8-97.3) | Ref | Ref | 501/510 (98.2, 96.7-99.1) | Ref | Ref |
| Nyakabwera | 1044/1070 (97.6, 96.5-98.3) | 1.72 (0.95-3.09) | 1.96 (1.05-3.60) * | 1046/1070 (97.8, 96.7-98.5) | 0.78 (0.34-1.64) | 0.80 (0.35-1.68) |
| Rubuga | 235/314 (74.8, 69.8-79.3) | 0.13 (0.08-0.21) *** | 0.13(0.07-0.23) *** | 304/314 (96.8, 94.2-98.3) | 0.55 (0.21-1.37) | 0.59 (0.23-1.49) |
| Kitoma | 361/430 (84.0, 80.2-87.1) | 0.22 (0.13-0.37) *** | 0.32(0.18-0.55) *** | 403/430 (93.7, 91.0-95.6) | 0.27 (0.12-0.56) *** | 0.29 (0.13-0.61) ** |
| Ruko | 226/359 (63.0, 57.8-67.8) | 0.07 (0.04-0.12) *** | 0.08 (0.05-0.14) *** | 326/359 (90.8, 87.4-93.4) | 0.18 (0.08-0.36) *** | 0.20 (0.09-0.41) *** |

**OR: Odds ratio, CI: Confidence interval, \*\*\*p<0.001, \*\*p<0.01, \*p<0.05**

## DISCUSSION

This community cross-section survey was conducted to assess the performance of three malaria diagnostic methods (RDTs, microscopy and qPCR) among community members (with or without symptoms of malaria) with a focus on asymptomatic individuals in areas where increasing levels of parasites with ART-R have been recently reported [42]. The study area is located near Rwanda and Uganda borders with human movements and where ART-R has been confirmed [9] Thus, this area is of high interest and it is being targeted as part of the response to ART-R with a broader focus on the Great Lakes Region of Africa. In this and other communities, recent studies have reported high prevalence of malaria among asymptomatic individuals and identified vulnerable groups (eg. males, school children, individuals with low socio-economic status and those living in poorly constructed houses) which need to be targeted by malaria interventions including intensified surveillance based on use of sensitive diagnostic methods [34,43]. In this study, the overall prevalence of malaria was higher by RDT compared to microscopy, and it was also higher among males, school children and individuals with fever history or fever at presentation. The prevalence of malaria infections was higher by RDTs followed by qPCR while microscopy had the lowest prevalence. The GMPDs of all species (*P. falciparum, P. malariae* and *P. ovale)* were higher by qPCR compared to microscopy. Using qPCR as the gold standard, RDTs had a higher sensitivity compared to microscopy while the specificity was higher for microscopy compared to RDTs, and the sensitivity of both RDTs and microscopy increased with increasing parasite density. These findings support the use of RDTs as a reliable parasite detection method for targeting community members, particularly asymptomatic individuals, and can potentially be utilized in the ongoing plans to develop a response to ART-R [9,42].

The prevalence of malaria infections was higher for RDTs while microscopy had the lowest prevalence. RDTs had the highest prevalence even for individuals of different age groups and sex. The high prevalence by RDTs could be due to the persistence of HRP2 antigens, which can remain detectable for up to four weeks after effective treatment, even after complete clearance of malaria parasites [42]. Higher prevalence of RDTs could also be caused by human errors during interpretation of results as some bands on test lines can be reported as present while the test is actually negative [44]. This means that some of the RDT positive results could actually be false positives, resulting high false positive results which has been associated with unwarranted prescription of antimalarials. History of antimalarial use within one to two weeks prior to testing may provisionally may help to identify individuals with false positive results as shown in this study that such individuals were more likely to be positive by RDTs and qPCR compared to microscopy, but cannot rule out failed clearance (recrudescence) or new infection.

The lower prevalence by microscopy compared to RDTs could be due to its low sensitivity particularly in low parasite density among asymptomatic individuals, quality of blood smears, and technical limitations. Previous studies have shown that low parasitemia below the limit of detection of microscopy is associated with an increasing rate of false negative results since a number of positive samples with low density infections are missed [45,46]. Quality and technical limitations including the optical condition of microscopes, skills of slide readers, smearing and slide staining quality have also been reported to affect microscopy results [47–49]. With poorly prepared smears or faulty microscopes, even appropriately-trained readers can potentially miss or misdiagnose malaria. Poorly trained microscopists can also contribute to this problem even if smears have been well prepared [47]. However, the study team used high quality reagents and experienced experts suggesting that these factors could not have potentially affected the results of this study. Thus, more studies will be needed to further tease out the poor performance of microscopy in similar study groups and areas of comparable transmission intensities.

Parasite densities were significantly higher for qPCR compared to microscopy, and this could be attributed to the low detection limit of qPCR compared to microscopy. Studies have shown that qPCR can identify as few as one parasite per microliter of blood, whereas microscopy typically requires a minimum threshold of around 50-100 parasites/μL) to ensure accurate identification [16,17]. Furthermore, parasitemia due to *P. falciparum* was higher compared to *P. ovale* spp and *P. malariae* with the highest *P. falciparum* parasitemia in under-fives. This could be due to differences in the biology of these malaria parasite species whereby *P. falciparum* is known to be highly pathogenic compared to other species, and this it is capable of rapid replication and the potential to evade the immune system leading to high parasitaemia [4,50]. The findings of this study are similar to what was reported in previous studies [51]. Low level of immunity among under-fives could be the main reason for higher parasitemia in this group, and this should always be considered when implementing case management strategies in under-fives [52].

While both RDTs and microscopy worked well, RDTs had a higher diagnostic accuracy than microscopy, and higher sensitivity but substantially lower specificity and PPV compared to microscopy. The specificity and PPV of RDTs were lower as expected, and it was most likely due to HRP2/3 protein residues that tend to persist even after the infection has been cleared with antimalarials [31,36]. RDTs were more sensitive than microscopy even at low parasitemia (<100 asexual parasites/μL), with the sensitivity of the two methods increasing as parasite density increased. The lower sensitivity of microscopy was potentially attributed to lower parasite density, as this study focused on community members whereby majority of the study participants (69.9%) were asymptomatic, with some of them carrying low-density infections below the detection limit of microscopy. The false positivity rate was higher with RDTs (12.2%) compared to microscopy (3.8%) suggesting that using RDTs particularly in community members made of mainly asymptomatic individuals should be properly assessed to avoid unwanted prescription of antimalarials, while the false negativity rate was higher with microscopy (25.0%) and lower with RDTs (6.8%). This agrees with the historical use of microscopy as a confirmatory test for malaria diagnosis due to its higher specificity [53]. However, as demonstrated from this study, RDTs, which are the current option for malaria testing at health facilities, had high sensitivity but low specificity, and microscopy which is the current gold standard, has high specificity but relatively lower sensitivity. Molecular detection by qPCR is highly sensitive and specific, but not feasible in clinical settings. More efforts need to be invested to determine the most ideal approach for malaria diagnosis in areas with heterogeneous malaria transmission. Such test will also be critical for supporting malaria surveillance within the ongoing elimination efforts and in responding to ART-R.

Parasite density, age, and sex were found to affect the sensitivity and specificity of RDTs and microscopy in various ways in this study. The sensitivity of both RDTs and microscopy increased with increasing parasite density and more false negatives were associated with low parasite density. Similar to what has been reported by others [45,54], the sensitivity of microscopy and RDT becomes very low below 100 parasites /µL or < 0.002% parasitaemia for RDTs and <50 parasites /µL or < 0.001% parasitaemia for microscopy. This implies that at low parasite densities, a considerable proportion of positive individuals may be missed by the tests, and this is of concern especially in areas targeting elimination as these are characterized with low level infections. The sensitivity of RDTs decreased with increasing age and this was similar to what was reported by others [55–57]. This could be explained by age-dependent immunity which develops following repeated exposure to infections, that may suppress parasites and result in low densities below detection threshold [56,58]. The specificity of RDTs increased with age, which is in agreement to what was previously reported by others [59–61], although other studies reported no age-specific trends[16,62]. The effect of age on specificity is thought to be influenced by the parasite density, which is related to the improvement of the immune system with age [59].

This study had two limitations. Firstly, a history of fever within the past two days and a history of antimalarial use within the previous seven days were based on self-reported information, increasing their potential recall bias. Participants or guardians of participants were the only source of information and the team had no means to ascertain their responses. However, the findings reported in this study are similar to what have been previously reported [23], suggesting that the responses reported in this study could potentially represent the actual status of fevers in the communities. Secondly, the study covered only one district of Kagera region where ART-R has recently been confirmed and the Ministry of Health is planning to implement a response strategy for ART-R. Thus, the findings from this study cannot be used to represent general performance of these three diagnostic methods in other areas of Mainland Tanzania. Despite these limitations, the finding of this study demonstrates higher performance of RDTs compared microscopy with qPCR as the reference method, suggesting that RDTs can be used as reliable methods for detection of malaria in communities with focus on areas with reported ART-R or ongoing malaria elimination strategies.

## CONCLUSION

This study revealed that RDTs were more sensitive and accurate but less specific compared to microscopy in detecting malaria parasites among community members, with a high proportion of asymptomatic individuals. The false positivity rate was higher with RDTs while the rate of false negative results was high with microscopy. The performances of both RDTs and microscopy were poor at very low parasite density (<100 parasite/μL), but increased with an increase in parasite density. The higher sensitivity and diagnostic accuracy of RDTs compared to microscopy supports the routine use of RDTs for case management and surveillance of asymptomatic malaria in this area with confirmed ART-R. Due to lower performance of microscopy particular among individual with low parasite density, RDTs usage in routine malaria diagnostic services should be prioritized, however, microscopy should be utilized for malaria confirmation purposes due to its high specificity. To ensure the high quality of malaria diagnosis results, the performance of RDTs and microscopy should be regularly monitored for supporting appropriate treatment of malaria infections with effective antimalarials as part of the strategies to fight ART-R

## Data Availability

All data produced in the present study are available upon reasonable request to the corresponding author

## Ethics approval and consent to participate

This CSS was part of the MSMT project whose protocol was reviewed and approved by the Medical Research Coordinating Committee (MRCC) of the National Institute for Medical Research (NIMR). Authorization to conduct the study was obtained from the President’s Office, Regional Administration and Local Government (PO-RALG), regional authorities, and the District Executive Directors. Information about the CSS was disseminated in the community through their village mobilization teams for two consecutive days preceding the survey. Before participating in the survey, verbal and written informed consent were sought and obtained from all participants or parents/guardians in the case of children.

## Availability of data and materials

The data used in this paper are available and can be obtained upon request from the corresponding author.

## Authors contribution

DSI developed the idea, supervised study implementation, data analysis, and interpretation of the results. FF, DPC, and MDS were involved in data collection, analysis, and results interpretation. RBM drafted the manuscript and all authors revised the manuscript. DSI revised and finalized the manuscript and all authors read and approved the manuscript.

## Acknowledgments

The authors sincerely thank the participants for their willingness to participate in the CSS, consenting, and contributing to the study. They also extend their gratitude to the data collection and laboratory teams for their valuable contributions, including Ezekiel Malecela, Oswald Oscar, Ildephonce Mathias, Gerion Gaudin, Kusa Mchaina, Hussein Semboja, Sharifa Hassan, Salome Simba, Hatibu Athumani, Ambele Lyatinga, Honest Munishi, Anael Derrick Kimaro, Ally Idrissa and Amina Ibrahim. Special thanks to the finance, administrative, and logistic support teams at NIMR: Christopher Masaka, Millen Meena, Beatrice Mwampeta, Neema Manumbu, Arison Ekoni, Sadiki Yusuph, John Fundi, Fred Mashanda, Amir Tununu and Andrew Kimboi. The support from the management of NIMR, NMCP and PO-RALG was critical to the success of this CSS, and it is therefore appreciated. The team extends gratitude for the technical and logistics support from partners at Brown University, the University of North Carolina at Chapel Hill, the CDC Foundation and the Bill and Melinda Gates Foundation team.

## Abbreviations and acronyms

ACT: Artemisinin-based combination therapy
AL: Artemether-Lumefantrine
aOR: Adjusted odds ratio
ART-R: Artemisinin partial resistance
CI: Confidence interval
CDC: Centers for Disease Control and Prevention
cOR: Crude odds ratios
CSS: Cross-sectional survey
DBS: Dried blood spots
DMFP: District malaria focal person
GMPD: Geometric Mean Parasite Density
IDs: Identification numbers
IPTp: Intermittent preventive treatment
IQR: Interquartile range
IRS: Indoor residual spraying
ITN: Insecticides treated nets
LSM: Larval source management
MRCC: Medical Research Coordinating Committee
MSMT: Molecular surveillance of malaria in Tanzania.
NIMR: National Institute for Medical Research
NMCP: National Malaria Control Program
ODK: Open Data Kit software
ORs: Odds ratio
PCA: Principal component analysis
PO-RALG: President’s Office, Regional Administration and Local Government
RDTs: Rapid diagnostic tests
rRNA: Ribosomal Ribonucleic Acid
SMPS: School-children Malaria Parasitological Survey
SOPs: Standard Operating Procedures
WHO: World Health Organization

